# Predicting PPE use, post-traumatic stress, and physical symptoms during the early weeks of COVID-19 lockdowns in the USA

**DOI:** 10.1101/2020.07.27.20162057

**Authors:** William H. O’Brien, Shan Wang, Aniko Viktoria Varga, Huanzhen Xu, Tracy E. Sims, Kristin A. Horan, Chung Xiann Lim

## Abstract

The COVID-19 pandemic combined with inconsistent governmental and public health recommendations, media communications emphasizing threat, and widespread lockdowns created a complex psychological environment for Americans. In this study, 450 MTurk workers completed measures of (a) risk for COVID-19, (b) perceived vulnerability to disease, (c) intolerance of uncertainty, (d) mindfulness, (e) COVID-19 preventive health behaviors, (f) post-Trauma symptoms, and (g) stress related physical symptoms. The surveys were completed between April 9, 2020 and April 18, 2020 which is a period that corresponded to the first 2-3 weeks of lockdown for most participants.

A substantial number of participants reported a reduction employment status and 69% were in self-isolation. The participants reported a high degree of perceived vulnerability with 68% indicating they felt there was a 50/50 chance or greater they would contract COVID-19. Mask wearing was variable: 16% “not at all,” 20% “some of the time,” 42% “a good part of the time,” and 26% “most of the time.” Using clinical cutoff on the post-trauma scale, 70% of the sample would be considered to have symptoms consistent with PTSD. The mean level of physical symptoms was significantly (*p* < .001) and substantially higher (d = 1.46) than norms.

PPE use was positively associated with level of education and mindfulness nonreactivity and negatively associated with age, having a current medical condition, and mindfulness nonjudgment. Post trauma and physical health symptoms were strongly predicted by susceptibility variables and intolerance of uncertainty.

**Highlights:**

- COVID-19 created a complex psychological environment for Americans due to threat exposure combined with contradictory communications from government and media.
- In a survey of 450 Americans, 68% reported that there was a 50/50 chance of greater they would contract COVID-19 and 70% of participants reported symptoms that met criteria for PTSD.
- Mask wearing was variable with only 26% reporting use “most of the time.”
- Participants who reported: older age, having one or mode medical conditions, less educational attainment, and less judgmental attitudes about their own thinking reported lower PPE use.
- Intolerance of uncertainty and perceived susceptibility were associated with higher PTSD symptoms.
- Mindfulness awareness and being judgmental attitudes about thinking were associated with lower PTSD symptoms.

## The COVID-19 Social and Psychological Context in Early 2020

The COVID-19 pandemic brought about rapid and dramatic changes in organizational, social, family, and individual behavior. After China confirmed the presence of COVID-19 in Wuhan on December 31, 2019 (WHO, 2020), United States citizens were initially assured that COVID-19 would be confined to China (Poynter Institute, 2020). A national emergency declaration was on March 13, 2020. At the same time and for weeks thereafter, contradictory communications and recommendations were widespread among government leaders and policymakers. These contradictory messages focused on key COVID-19 prevention and containment issues such as: mask wearing, hydrocholoroquine use, social distancing, COVID-19 testing, and the need for lockdowns.

Through January and February, the CDC communicated that COVID-19 risk was minimal and that no specific preventive actions were needed other than avoiding travel to and from affected countries (CDC, 2020). It is now recognized that during this time many undiagnosed cases of COVID-19 were present and spreading in the USA (Deng et al., 2020; Shear, Goodnaugh, Kaplan, Fink, Thomas, & Weiland, 2020). In March and April, the CDC issued recommendations that more clearly articulated the degree of COVID-19 risk and generated preventive health recommendations that included lockdowns, social distancing, testing, contact tracing, and personal protective equipment (PPE) use (CDC, 2020).

While governmental communications and recommendations where contradictory, news media sites were emphasizing threat. USA infection rates and death counts were prominently featured along with extensive reporting of the scarcity of medical resources, a possible economic catastrophe, and the social impact of COVID-19 (Ioannidis, 2020). Oftentimes, the headlines were marked by fear-evoking terms such as “apocalyptic.” Our search of Newspaper.com on June 18, 2020, yielded 80,5873 print stories on “Covid-19 and Death” compared to 3,974 stories on “Covid-19 and survival.”

By mid to late March, the magnitude of health threat posed by COVID-19 prompted declarations of states of emergency, quarantines, closing of businesses and schools, and social distancing guidelines. By late April, most USA states had implemented COVID-19 quarantine and social distancing procedures. As such, most persons in the USA were confined to homes and residences, no longer working or attending school, and prohibited from participating in many reinforcing social and recreational activities.

A similar set of circumstances occurred with the H1N1 pandemic in 2009. Taha and colleagues (Taha, Matheson, Anisman, 2014a; 2014b; Taha, Matheson, Cronin, & Anisman, 2014) explored the interplay between governmental responses, public health official communications, and media reporting. Their research indicated that inconsistencies across these entities likely contributed to higher levels of anxiety, mistrust, and poor adoption of preventive health behaviors. In a prescient cautionary statement, Taha et al, (2014a) noted “Although the risk for contracting H1N1 has subsided, virologists have suggested another pandemic will occur again…that will increase transmission and/or lethality. This means that the public may again have to contend with a health threat with largely uncertain consequences.” (p. 149). They added in a different article “clearly governmental agencies and media outlets need to ensure that they have one voice, so that the delicate balance between conveying the potential severity of a pandemic on the one hand, and preventing panic on the other, can be negotiated” (Taha et al, 2014, p. 603). Taha and colleagues could not have envisioned the degree of inconsistency that was presented to the American public during the COVID-19 pandemic or that key leaders would recommend and model *noncompliance* with public health recommendations. Nonetheless, their cautionary statements are particularly relevant to understanding the current COVID-19 psychological context experienced by Americans.

Perceived uncertainty, unpredictability, and inconsistency of information regarding effective prevention actions under conditions of threat exposure have been reliably linked to psychological distress (Carleton, 2016; Contrada & Baum, 2011). The introduction of significant lifestyle restrictions associated with quarantines and lockdown would intensify this distress due to a reduction in social, recreational, financial, and occupational reinforcement (Ahmed, Ahmed, Aibao, Hanbin, Siyu, & Ahmad, 2020). The combined effects of information and recommendation inconsistency, threat magnification via media reporting, and significant lifestyle changes created a complex and likely harmful psychological environment for many Americans.

## A Functional Contextual Behavioral Approach to Preventive Health Behavior

Exposure to verbal or visual health threat information evokes psychophysiological activation that is experienced as an aversive state (Contrada & Baum, 2011; Hyde, Ryan, & Waters, 2019). In the health psychology literature, this aversive state has sometimes been labeled “perceived susceptibility” and it has been associated with engagement in preventive health behaviors (Brewer, Gibbons, Gerrard, McCaul, & Weinstein, 2007; O’Brien, Mumby, & VanEgeren, 1995; Sheeran, Harris, & Epton, 2014). Under conditions of health threat and the concomitant increase in perceived susceptibility, avoidance and escape behaviors will be generated. Avoidance and escape behaviors that lead to a reduction in perceived susceptibility are more apt to be repeated via negative reinforcement.

Avoidance and escape behaviors can be cognitive and/or behavioral - more specifically overt-motor (Hayes, Wilson, Gifford, Follette, & Strosahl, 1996). Cognitive avoidance and escape behaviors can include information seeking, problem solving, rumination, and worry. These cognitive responses are labelled experiential avoidance because they provide a means for diverting attention away from aversive imagery or emotional experiencing of threat information (Boulanger, Hayes, & Pisterello, 2010). Cognitive avoidance and escape responses can be adaptive or workable to the extent that they promote engaging in behaviors that lead to a better quality of life or thriving (e.g., generating effective solutions). They can also be maladaptive and unworkable to the extent that they lead to a poorer quality of life and lack of thriving (e.g., persistent ruminating and immobilization).

Overt-motor avoidance and escape responding under conditions of general threat and health threat has been well-researched in the psychological literature (e.g., Dymond, Schlund, Roche, De Houwer, & Freegard, 2012; Tannenbaum, Hepler, Zimmerman, Saul, Jacobs, Wilson, & Albarracín, 2015). Related to health threat information, adaptive and workable overt-motor avoidance and escape behaviors include engaging in effective preventive health actions, seeking social support, and engaging in meaningful activity. Alternatively, maladaptive behaviors include those that reduce quality of life and thriving such as noncompliance with preventive actions and substance abuse.

Applied to COVID-19, the functional contextual behavioral perspective supports a prediction that exposure to health threat information would prompt an increase in perceived susceptibility and the concomitant motivation to engage in avoidance and escape responses. Cognitive and overt-motor avoidance and escape responses that bring about a reduction in objective risk and simultaneously reduce the intensity, duration, and/or frequency of perceived susceptibility are more workable than responses that do not reduce objective risk (Presti, McHugh, Gloster, Karekla, & Hayes, 2020). Adaptive behaviors such as PPE and social distancing can reduce the objective risk of COVID-19 transmission and infection (e.g., Cook, 2020).

## Traits Associated with Health Threat Reactivity

A general self-view of vulnerability to disease has also been identified and found have cross-situational and cross-temporal consistency (Duncan, Shaller, & Park, 2009). Persons with higher levels of general perceived vulnerability to disease are more apt to show higher levels of distress in response to health threat information. Intolerance of uncertainty is a trait that has also been associated with health threat reactivity. Carleton (2016b) defined intolerance of uncertainty as “an individual’s dispositional incapacity to endure the aversive response triggered by the perceived absence of salient, key, or sufficient information, and sustained by the associated perception of uncertainty” (p. 31). Intolerance of uncertainty has been associated with heightened psychophysiological reactivity and conditioning to threat cues (Morriss, Saldarini, & van Reekum, 2019) and impairment of safety learning (Wake, van Reekum, Dodd, & Morriss, 2020).

Trait mindfulness is characterized by intentional awareness and present-moment focus where external information and internal experiences are acknowledged and viewed from a nonreactive and nonjudgmental perspective (Shapiro, Carlson, & Freedman, 2006). Applied to COVID-19, one would expect that higher levels of mindfulness would be associated with lower levels of distress. It is less clear how mindfulness would be associated with preventive health behaviors. Some researchers have linked mindfulness to higher levels of safety behavior and preventive health actions (O’Brien, Horan, et al, 2019). The interpretation of these findings is that mindfulness may moderate the influence of thoughts that interfere with the adoption and maintenance of preventive health actions. However, to the extent that mindfulness is associated with lower levels of distress (including perceived susceptibility) under threat conditions, mindfulness could be associated with lower levels of preventive health behavior.

## Summary

The genesis of this investigation occurred during the emergence of the COVID-19 pandemic in China. The social and psychological context was unprecedented. A highly transmissible, poorly understood, lethal virus was rapidly spreading across the United States. Media reporting of hospitalizations and deaths were extensive and dramatic. Susceptibility information and recommendations on how to reduce risk were fractionated. Ultimately, self-isolation and quarantine were developed and enforced with varying levels of support from leadership. With the self-isolation and quarantines, large numbers of people experienced a significant loss of opportunities to engage in reinforcing occupational, social, recreational, and physical activities.

This project was developed as an effort to document levels of distress experienced by persons in the USA during the early stages of lockdown. We also sought to evaluate the relationships among objective risk factors for COVID-19, perceived susceptibility for COVID-19, trait variables that would be associated with distress (perceived vulnerability to disease, intolerance of uncertainty), mindfulness, and reported use of preventive health behaviors (PPE use). We expected that the combination of chronic exposure to health threat combined with a contradictory information environment be manifested in inconsistent use of preventive health behaviors, higher levels of psychological distress, and higher levels of stress-related physical symptoms. We also expected that perceived vulnerability and intolerance of uncertainty would be associated with higher levels of perceived susceptibility to COVID-19 which in turn would predict engagement in preventive health behaviors. Finally, we expected that mindfulness would be associated with lower levels of distress and physical symptom reporting and higher levels of engagement in preventive health behaviors.

## Methods

### Participants

This project was approved by the Bowling Green State University Institutional Review Board on March 30, 2020 (#1562479-4). Amazon Mechanical Turk Workers were enrolled through CloudResearch (Litman, Robinson, & Abberbock, 2016). Six hundred thirty five participants initially responded to the survey from April 9, 2020 to April 18, 2020. Of these, 116 started the survey but discontinued quickly (all 116 participants completed less than 20% of the survey items and 105 completed less than 10%). Data quality for the remaining 519 participants was examined. Participant data was deleted if any of the following were detected: (a) less than 75% of items completed (n = 15), duplicate IP address (n = 26), (b) failing 2 of three attention check items (n = 27), or (c) an unusually long time (231 minutes) to complete the survey (n = 1). This resulted in a final total sample of 450 participants.

The demographic characteristics of participants are summarized in Table 1 and the geographic distribution is depicted in Figure 1. The mean age was 36.68 (range 18 – 76) and 37% of the participants were female. The sample was predominantly Caucasian (55.3%) followed by Black/African American (22.9%) and Asian (4.4%). About half (55.6%) participants reported obtaining a Bachelor’s degree and 26.5 % reported obtaining a Master’s degree or higher. These statistics indicate that relative to census data, there was a higher proportion of males, Black/African Americans, and persons with higher educational attainment in this sample (retrieved from www.census.gov). Sixty-six (15%) of the participants reported having one or more medical condition. The more commonly reported conditions were cardiovascular disease, diabetes, lung diseases (COPD, asthma), and autoimmune diseases.

**Table 1.** Sample demographics

| Variable |  | M (SD) or N (%) |
| --- | --- | --- |
| Age |  | 36.68 (11.27) |
| Sex |  | 1<br>70 (37.8%) Female |
| Employment (prior to COVID-19) |  |  |
|  | Employed (1-24 hrs./wk.) | 72 (16%) |
|  | Employed (24-39 hrs./wk.) | 110 (24.4%) |
|  | Employed (40 or more hrs./wk.) | 240 (53.3%) |
|  | Unemployed (looking for work) | 10 (2.2%) |
|  | Unemployed (NOT looking for work) | 11 (2.4%) |
|  | Retired | 3 (.7%) |
|  | Disabled (unable to work) | 4 (.9%) |
| Employment (after COVID-19) |  |  |
|  | Employed (1-24 hrs./wk.) | 98 (21.8%) |
|  | Employed (24-39 hrs./wk.) | 134 (29.8%) |
|  | Employed (40 or more hrs./wk.) | 164 (36.4%) |
|  | Unemployed (looking for work) | 21 (4.7%) |
|  | Unemployed (NOT looking for work) | 23 (5.1%) |
|  | Retired | 3 (.7%) |
|  | Disabled (unable to work) | 7 (1.6%) |
| Marital status |  |  |
|  | Single | 88 (19.6%) |
|  | Cohabiting (not married) | 21 (4.7%) |
|  | Long term relationship (not married or cohabitating) | 12 (2.7%) |
|  | Married | 309 (68.7%) |
|  | Divorced | 17 (3.8%) |
|  | Widowed | 3 (.7%) |
| Children |  | 306 (68%) with children |
| Education |  |  |
|  | High school (or GED) | 18 (4.0%) |
|  | Some college (no degree) | 37 (8.2%) |
|  | Associate's degree | 26 (5.8%) |
|  | Bachelor's degree | 250 (55.6%) |
|  | Master's degree or higher | 119 (26.5%) |
| Race |  |  |
|  | Hispanic/Latino | 49 (10.9%) |
|  | Caucasian | 249 (55.3%) |
|  | Black/African American | 103 (22.9%) |
|  | Asian | 20 (4.4%) |
|  | Native Hawaiian/Pacific Islander | 2 (.4%) |
|  | American Indian/Alaska Native | 6 (1.3%) |
|  | Two or more races | 13 (2.9%) |
|  | Choose not to identify | 8 (1.8%) |
| Religion |  |  |
|  | Protestant | 84 (18.7%) |
|  | Roman Catholic | 243 (54%) |
|  | Mormon | 2 (.4%) |
|  | Jewish | 15 (3.3%) |
|  | Muslim | 10 (2.2%) |
|  | Buddhist | 8 (1.8%) |
|  | Taoist | 4 (.9%) |
|  | Hindu | 7 (1.6%) |
|  | Agnostic | 28 (6.2%) |
|  | Atheist | 25 (5.6%) |
|  | Nothing in particular | 29 (6.4%) |
|  |  | 66 (15%) |
| Medical Condition |  |  |
|  | Yes | 66 (15%) |
|  | No | 383 (85%) |

**Figure 1.**
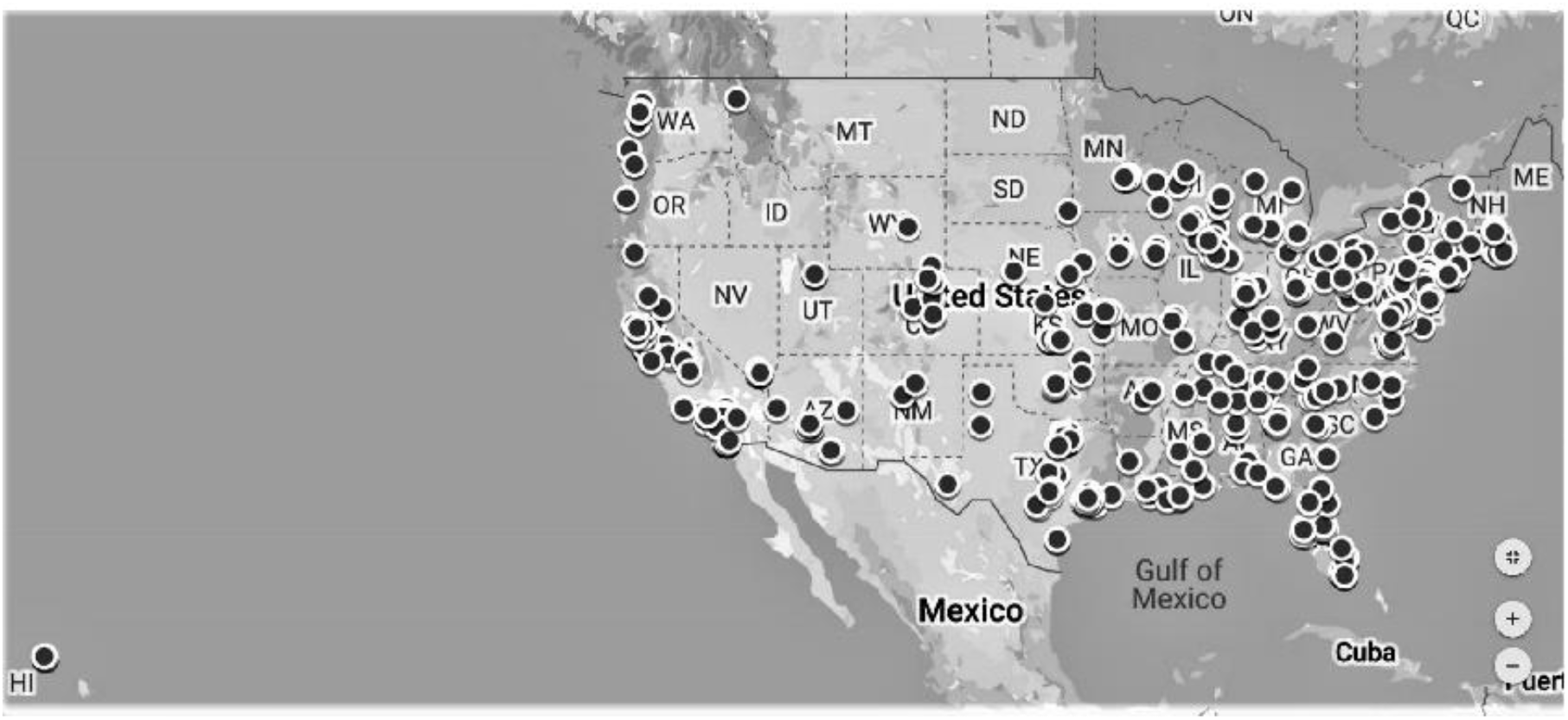
Geographic Distribution of Participants.

### Measures

#### Demographics

Participants completed items that provided information about basic demographic characteristics: age, sex, race, ethnicity, marital status, educational attainment, number of children, living circumstances. They also provided information about employment (job type, hours per week, changes in job since COVID-19). Participants reported whether they were experiencing any medical conditions and listed medications they were taking.

#### Self-isolation, quarantine, and COVID-19 risk exposure

Participants reported whether they were engaged in self-isolation or quarantine. They also reported the number of times they leave their residence in a typical day and how many times they left their residence in the past month. Finally, participants reported whether they had been in any setting in the last 14 days that they would consider to be risky for contracting COVID-19. The rating ranged from 1 (“extremely low”) to 5 (“extremely high”).

#### Perceived vulnerability to disease (PVD)

The PVD is a 15-item scale designed to measure general perceptions of risk for illness (Duncan, Schaller, & Park, 2009). Our analysis of the internal consistency of the subscales recommended by Duncan et al (2009) revealed suboptimal Cronbach’s alphas (α = .57 for infectibility and α = .54 for germ aversion). We therefore conducted an exploratory factor analysis with an oblimin rotation to identify subscales that fit the current sample. Results indicated that there were three factors that accounted for 56.44% of the variance (factor 1 = 30.96%, factor 2 = 13.87%, factor 3 = 11.62%).

Item loadings were evaluated. If an item loaded more than .40 on a single factor and there was at least a .20 difference in loadings on any other factor (Hinkin, 1998), it was retained. If an item loaded at .32 or higher on two or more factors it was considered cross-loaded and removed. Four items were removed due to low factor loadings or cross loadings. The remaining 11 items all had high loadings (> .69) one of the three factors. An examination of items indicated that factor one (4 items) was measuring *general perceived vulnerability to disease* (e.g., “In general, I am very susceptible to colds, flu, and other infectious diseases”), factor 2 (4 items) was measuring *perceived lack of immunity* (e.g., “my immune system protects me from most illnesses that other people get” - reversed coded) and factor 3 (3 items) was measuring *germ aversion* (e.g., “I don’t like to write with a pencil someone else has obviously chewed on”). The correlations among the three factors were low indicating limited overlap in constructs: general vulnerability and lack of immunity, r = −.29; general vulnerability and germ aversion, r = −.004; and lack of immunity and germ aversion, r = −.02. The internal consistencies of the three subscales were: susceptibility (α = .87), lack of immunity (α = .71), and germ aversion (α = .56). The low reliability of the germ aversion factor could not be improved by adding or deleting items. For each subscale higher scores indicated more general vulnerability, less perceived immunity, and more germ aversion.

#### Perceived vulnerability to COVID-19

A single item was constructed to assess perceived vulnerability to COVID-19. The item was worded “How likely is it that you will contract COVID-19?” Response options ranged from “no chance” to “certain” using a five-point scale.

#### Mindfulness - Five Facet Mindfulness Questionnaire (FFMQ)

The FFMQ is a scale designed to measure mindfulness in daily life (Baer, Smith, Hopkins, Krietemeyer, & Toney, 2006). The measure used in this study was the 24-item version that generated five subscales: awareness, observe, describe, nonjudgment, and nonreactivity. Participants rated items using a 5-point Likert-type scale ranging from 1 (*never* or *very rarely true*) to 5 (*very often* or *always true*). The internal consistency across all FFMQ items was Cronbach’s alpha = .79. The internal consistencies of each subscale were: Awareness (α = .87), observe (α = .72), describe (α = .61), nonjudge (α = .80), and nonreactivity (α = .80). Total scores were calculated for each subscale with higher scores indicating higher levels of mindfulness.

#### Intolerance of uncertainty

The 12 item version of the intolerance of uncertainty scale (IUS, Carleton, Norton, Asmundson, 2007; Carleton, Collimore, & Asmundson, 2010) was used to assess psychological distress associated with ambiguity and unpredictability (e.g., “when I am uncertain, I can’t function very well”). The IUS has been evaluated in student and community samples (Hale et al., 2016) with results indicating that it measures inhibitory anxiety and prospective anxiety. The internal consistency of the IUS was very good (Cronbach’s α = .91). A total score was calculated with higher scores indicating higher levels of intolerance of uncertainty.

#### Preventive actions taken scale (PATS)

The PATS was developed in late January 2020 based on recommendations generated by then-available COVID-19 research findings. The original 12-item measure assessed the extent to which participants in a China community survey reported engaging in behaviors to prevent COVID-19 infection at two levels, i.e. a recommended level (e.g. wearing mask outside of home) and an excessive level (e.g. wearing mask everywhere).

We modified the PATS by taking out two items that were not as relevant to COVID-19 transmission in the USA (eating wild animal, eating any meat). We also eliminated two redundant items measuring mask and glove wearing. The remaining 8 items were factor analyzed using an oblimin rotation to identify subscales and item adequacy. The factor analysis yielded two factors that cumulatively accounted for 58.14% variance (factor 1 = 32.81%, factor 2 = 25.32%).

Item loadings were examined using the same procedures described above No items were removed based on the factor analysis. The remaining 8 items all had high loadings (> .68) on one of the two factors. Factor one (5 items) measured *avoiding public settings and contact with people* (e.g., “I avoid public events and crowded places”), factor 2 (3 items) measured *PPE use* (“I wear a mask everywhere”). The correlation among the two factors was low, r = .08. The internal consistencies of the two subscales were good: avoiding public settings and contact with people, Cronbach’s α = .76; PPE, Cronbach’s α = .77. For both subscales, higher scores indicated more engagement in preventive behavior.

#### Psychological symptoms

The Impact of Events Scale – Revised (IES-R) was used to measure post-traumatic stress symptoms (Weiss & Marmar, 1997). The IES-R has been extensively used as a measure of post-traumatic stress symptoms in community and clinical samples (e.g., Beck, et al., 2008). The scale contains 22 items (e.g. “I thought about it when I didn’t mean to”) that were responded to on a five-point scale that ranged from 0 (“not at all”) to 4 (“extremely”). Participants were instructed to indicate how distressing each difficulty has been for the past 7 days with respect to the Coronavirus situation. The internal consistency of the scale was high (Cronbach’s α = .97). A total IES-R score was calculated with higher scores indicating higher levels of symptoms.

#### Physical symptoms

The Patient Health Questionnaire (PHQ-15, Kroenke, Spitzer, & Williams, 2002)) was used to measure physical symptoms associated with stress. For each item (e.g., “dizziness”), participants rated the degree of being bothered in the prior month using a 3-point Likert scale that ranged from 0 (“not bothered at all”) to 2 (“bothered a lot”). The internal consistency of the PHQ-15 in this study was high (Cronbach’s α = .92). A total score was calculated with higher scores indicating higher levels of physical symptoms.

### Procedure

An announcement was placed on Amazon Mechanical Turk on April 9, 2020. The announcement read “The COVID-19 situation is creating worldwide challenges. In this survey study, university researchers hope to gain important useful information about how people are reacting to COVID-19 and coping with COVID-19. The survey is intended to be taken by individuals who are at least 18 years old who reside in the United States. The survey should take around 20 minutes to complete…You will receive $1.00 for completing the survey.” Interested participants were then directed to the informed consent form which was linked to the survey. The survey contained 3 attention check items and 3 captcha items.

## Results

### Descriptive Statistics

Analyses of frequencies and measures of central tendency were conducted to characterize participant reports of employment and living circumstances, preventive actions, COVID-19 risk exposure, perceived vulnerability, psychological distress, and mindfulness.

#### Employment

As shown in Table 1, the numbers of unemployed and less than half-time employed (1-24 hours) doubled while the number of full-time workers decreased by 36% from pre to post COVID-19.

#### Self-Isolation and quarantine

Self-isolation was reported by 69% of the participants. The average number of days in self-isolation was 20 (*sd* = 14.13). A total of 31% of the participants reported “I am currently in quarantine” to the question “have you been ordered to quarantine,” 25% responded “yes and finished,” 42% responded “no,” and 2% responded “not yet, but will be due to travel plans.”

#### COVID-19 risk exposure

Forty percent (n = 180) of the participants reported being in a setting that exposed them to COVID-19 risk in the past 14 days. Of those reporting risk exposure, 37% (n = 66) rated the level of risk to be “extremely low” or “slight,” 29% (n = 53) rated the level of risk to be “moderate,” and 34% (n = 61) rated the level of risk to be “high” or “extremely high.”

#### Perceived vulnerability to disease and COVID-19

The three subscales yielded the following means and standard deviations: general perceived vulnerability (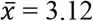, *sd* = 1.02), lack of immunity (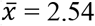, *sd* = .79), and germ aversion (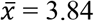, *sd* = .72). The differences among means was significant with a very large effect size (*F* (2, 898) = 234.68, *p* < .001; *η2* = .34). Follow-up pairwise comparisons were all significant (*p* < .001). These measures were therefore considered separately in further analysis.

Ratings of perceived risk of contracting COVID-19 indicated that 4% (n = 19) of the participants reported that they felt “certain” they would contract COVID-19, 27% (n = 121) reported they felt it was “likely,” 29% (n = 131) reported they felt there was a 50/50 likelihood, 29% (n = 130) reported they felt it was “unlikely,” and 10% (n = 47) reported they felt there was “no chance.”

#### Intolerance of Uncertainty

The mean score for the IUS was 39.81 (*sd* = 9.88). This mean was substantially higher than those reported by Carelton et al. (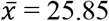, *sd* = 9.45, 2007) who collected data using an undergraduate sample. There are no comparison norms that match an Mturk sample.

#### Mindfulness

The means for the five FFMQ subscales were: Awareness (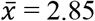, *sd* = 1.03), Describe (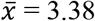, *sd* = .67), Observe (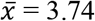, *sd* = .72), Nonreactivity (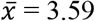, *sd* = .76), and Nonjudgement (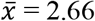, *sd* = .85). USA norms for the FFMQ-24 have not been published.

#### Preventive actions

The means for *PPE use* and *avoiding public settings and contact with people* were, respectively 2.70 (*sd* = .82) and 3.19 (*sd* = .60). The difference between the two ratings was significant with a large effect size (*F* (1, 449) = 112.48, *p* < .001; *η2* = .20) indicating that PPE use was a less frequently used preventive action relative to avoidance of public spaces and contact with others.

An item level analysis of the PPE use indicated that 16% of the participants reported that wearing a mask “does not apply to me at all/none of the time,” 20% reported “applies to me some degree or some of the time,” 42% reported “applies to me a considerable degree or a good part of the time,” and 26% reported “applies to me very much or most of the time.” A similar level of endorsement was observed for wearing gloves and precautionary purchases (respectively: 21%, 15%, 45%, 19%; 10%, 27%, 42%, 21%).

#### Psychological Distress and Physical Symptoms

The average IES-R total was (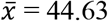, *sd* = 23.50). This average score is significantly higher (*t* (630) = 5.81, *p* < .0001; *d* = .53) than the mean value reported by Beck et al (2008) who measured a sample of persons who had experienced serious motor vehicle accidents (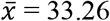, *sd* = 18.98, n = 182). Using clinical cutoffs (.91 sensitivity, .82 specificity) reported by Creamer, Bell, and Failla (2002), 70% of the participants in the current sample would be classified as reporting symptom levels that are consistent with PTSD.

The average PHQ-15 total was (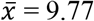, *sd* = 7.12), which is substantially and significantly (*d* = 1.46; *t* (5479) = 17.52, *p* < .00001) higher than the normative mean from a German sample (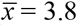, *sd* = 4.1) reported by Kocalevent, Hinz, and Brähler (2013).

### Predicting PPE use

Hierarchical regression analyses were used to predict PPE use. We did not conduct regression analyses for avoiding public settings and contact with people because, at the time of this study, most participants were under quarantine and self-isolation orders.

In the first step (model 1), demographic and risk variables (age, sex, education, current medical condition) were entered as predictors. In the second step (model 2), we added the trait measures of vulnerability to disease (general vulnerability, lack of immunity, germ aversion), specific perceived susceptibility to COVID-19, and intolerance of uncertainty. In the third step (model 3) we added the five FFMQ subscale scores. A Bonferroni correction was used to correct for multiple t-tests of regression coefficients. The Bonferroni correction yielded a p < .004 as the threshold needed for classifying a result as significant.

All three models were significant (see Table 2). The addition of variables in models 2 and 3 were associated with significant increases in R2. An examination of the contribution of individual variables indicated that reporting a medical condition was significantly associated with *less* PPE use and education level was associated with significantly more PPE use. None of the other demographic variables accounted for significant proportion of the variance although there was a trend for increases in age to be associated with less PPE use. None of the trait vulnerability measures, nor perceived susceptibility to COVID-19, nor intolerance of uncertainty significantly predicted PPE use. The FFMQ nonreactivity subscale was significantly positively associated with PPE use while the FFMQ nonjudgement subscale was significantly negatively associated with PPE use.

**Table 2.** Predicting PPE use

| *Predictor* | $\beta$ | *t* | *p* |
| --- | --- | --- | --- |
| Age | -.10 | -2.61 | .009 |
| Sex | .09 | 2.34 | .020 |
| Medical condition | -.18 | -4.41 | <.001* |
| Education | .13 | 2.98 | .003* |
| $R^2 = .21, F(4, 442) = 29.96, p < .001$ | | | |
| General Perceived Vulnerability | .13 | 2.33 | .020 |
| Perceived Poor Immunity | .09 | 1.69 | .093 |
| Germ Aversion | -.03 | -.60 | .551 |
| Susceptibility to COVID-19 | -.07 | -1.71 | .083 |
| Intolerance of Uncertainty | .11 | 1.93 | .055 |
| $Change\ in\ R^2 = .14, F(5, 437) = 18.35, p < .001; Total\ R^2 = .35, F(9, 437) = 26.12, p < .001$ | | | |
| FFMQ Aware | -.07 | -1.07 | .283 |
| FFMQ Describe | .02 | .44 | .661 |
| FFMQ Observe | -.03 | -.71 | .476 |
| FFMQ Nonreact | .17 | 3.50 | .001* |
| FFMQ Nonjudge | -.27 | 4.42 | <.001* |
| $Change\ in\ R^2 = .06, F(5, 432) = 9.37, p < .001; Total\ R^2 = .41, F(14, 432) = 21.75, p < .001$ | | | |

*Note.* FFMQ = Five Factor Mindfulness Questionnaire – 24, \* indicates significance after Bonferroni correction ( $p = .004$ ).

### Predicting Psychological Distress and Physical Symptoms

#### Post-Trauma Stress Symptoms

Hierarchical regression analyses were used to predict IES scores. The same variables that were used to predict PPE were used in these analyses. We added the self-isolation variable as another predictor variable to evaluate the possible effects of a reduction in access to reinforcing activities.

For the IES score, all three models were significant (see Table 3). The addition of variables in models 2 and 3 were associated with significant increases in R2. None of the demographic variables were significantly associated with the IES total. Increases in general perceived vulnerability and intolerance of uncertainty were both significantly associated with higher IES scores. Perceived lack of immunity (trend, but not significant after Bonferonni correction) and germ aversion were inversely associated with the IES score. The FFMQ awareness and nonjudgment subscales were significantly, positively, and nontrivially associated with lower IES scores.

**Table 3.** Predicting post trauma stress symptoms using the Impact of Experiences Scale

| *Predictor* | $\beta$ | $t$ | $p$ |
| --- | --- | --- | --- |
| Age | -.02 | -1.08 | .282 |
| Sex | .03 | 1.45 | .147 |
| Medical condition | -.05 | -2.14 | .033 |
| Education | .02 | .59 | .552 |
| $R^2 = .21, F(4, 442) = 29.07, p < .001$ | | | |
| General Perceived Vulnerability | .18 | 5.69 | <.001* |
| Perceived Poor Immunity | -.09 | -2.79 | .005 |
| Germ Aversion | -.10 | -3.84 | <.001* |
| Susceptibility to COVID-19 | -.01 | -.20 | .844 |
| Intolerance of Uncertainty | .31 | 9.74 | <.001* |
| $Change\ in\ R^2 = .53, F(5, 437) = 180.19, p < .001; Total\ R^2 = .74, F(9, 437) = 139.21, p < .001$ | | | |
| FFMQ Aware | -.20 | -5.24 | <.001* |
| FFMQ Describe | -.06 | -1.96 | .051 |
| FFMQ Observe | .03 | .97 | .332 |
| FFMQ Nonreact | .03 | 1.00 | .318 |
| FFMQ Nonjudge | -.21 | -6.04 | <.001* |
| $Change\ in\ R^2 = .06, F(5, 432) = 25.45, p < .001; Total\ R^2 = .80, F(14, 432) = 123.62, p < .001$ | | | |

*Note.* FFMQ = Five Factor Mindfulness Questionnaire – 24, \* indicates significance after Bonferroni correction ( $p = .004$ ).

#### Physical symptoms

For the PHQ-15 score, all three models were significant (see Table 4). The addition of variables in models 2 and 3 were associated with significant increases in R2. None of the demographic variables were significantly associated with the PHQ-15 total. General perceived vulnerability, perceived susceptibility to COVID-19, and intolerance of uncertainty were all significantly associated with increases in PHQ-15 scores. Perceived lack of immunity was inversely associated with PHQ-15 scores with significance levels failing to meet the Bonferonni-corrected cutoff. There was also a trend for FFMQ awareness to be inversely associated with PHQ scores.

**Table 4.** Predicting physical symptoms using the Patient Health Questionnaire 15

| *Predictor* | $\beta$ | *t* | *p* |
| --- | --- | --- | --- |
| Age | -.01 | -.35 | .726 |
| Sex | .01 | .06 | .952 |
| Medical condition | -.06 | -1.66 | .099 |
| Education | .02 | .42 | .678 |
| $R^2 = .09, F(4, 442) = 11.22, p < .001$ | | | |
| General Perceived Vulnerability | .17 | 3.30 | .001* |
| Perceived Poor Immunity | -.15 | -2.81 | .005 |
| Germ Aversion | -.06 | -1.27 | .204 |
| Susceptibility to COVID-19 | .14 | 3.68 | <.001* |
| Intolerance of Uncertainty | .20 | 3.82 | <.001* |
| $Change\ in\ R^2 = .35, F(5, 437) = 54.65, p < .001; Total\ R^2 = .441, F(9, 437) = 38.38, p < .001$ | | | |
| FFMQ Aware | -.12 | -1.93 | .054 |
| FFMQ Describe | -.02 | -.47 | .640 |
| FFMQ Observe | -.02 | -.38 | .703 |
| FFMQ Nonreact | -.01 | -.11 | .913 |
| FFMQ Nonjudge | -.10 | -1.75 | .082 |
| $Change\ in\ R^2 = .02, F(5, 432) = 2.55, p < .027; Total\ R^2 = .80, F(14, 432) = 26.02, p < .001$ | | | |

*Note.* FFMQ = Five Factor Mindfulness Questionnaire – 24, \* indicates significance after Bonferroni correction ( $p = .004$ ).

## Discussion

The participants in this study reported substantial reductions in employment levels and most reported they were in self-isolation. Despite these isolation efforts, most participants reported the felt that they were likely exposed to COVID-19 and that the exposure constituted a moderate to high risk. A majority of participants report showed that they believed they would eventually contract COVID-19.

Most participants reported some PPE use but a sizable minority (36%) reported none to minimal PPE use. High levels of trauma symptoms were reported with 70% of the scores on the IES-R exceeding clinical cutoffs for PTSD. Physical symptom reporting was also very high.

These results indicate that this sample of adults reported loss of access to work and income, loss of access to reinforcing activity, and exposure to real and/or perceived COVID-19 risk. Given the social and psychological context of the measurement period, they were also being exposed to frequent and intense and contradictory messaging about danger and appropriate risk reduction strategies. Isolation, loss of reinforcement, intense exposure to threat information, and amplification of uncertainty have been associated with psychological distress. When combined, they would be a potent mix for creating widespread and substantial psychological distress. This was evident in the current sample where high levels of PTSD symptoms and physical health symptoms were reported.

Higher risk participants – older and those with current medical conditions, were *less* likely to report using PPE. We investigated whether these inverse correlations were due to older persons and those with medical conditions being more likely to self-isolate and avoid contact with others in which case they would have a diminished need for PPE use. We also explored the possibility that low income might interfere with mask and glove purchases. We thus regressed PPE on age, sex, medical condition, education, income, self-isolation, the number of times the participant left the home in the past month, and avoiding public settings/contact with other people. Results indicated that age and current medical condition continued to be significant and inverse (age, β = −.13, *t* = 2.96, *p* = .003; medical condition, β = −.17, *t* = 3.89, *p* < .001). Education and sex continued to independently and significantly predict more PPE use (education, β = .24, *t* = 5.62, *p* < .001; sex β = .13, *t* = 3.10, *p* = .002. Income was not significantly associated with PPE use (β = .04, *t* = .93, *p* = .353).

The negative relationship between medical condition and use of PPE could be due to other third variables such as a general lack of engagement in preventive health behaviors. This speculation is based on the possibility that the many of the reported medical conditions in this sample (e.g., diabetes, hypertension) could be linked to self-care behaviors. Thus, lower levels of general self-care would predict both medical condition and reduced PPE use.

Perceived susceptibility, intolerance of uncertainty, FFMQ-nonreactivity predicted more PPE use while FFMQ-nonjudgment was associated with less PPE use. The susceptibility findings are consistent with the preventive health behavior literature (Brewer et al., 2007). Intolerance of uncertainty when combined with susceptibility would be apt to create a higher level of motivation to engage in preventive health behavior from a functional contextual perspective. Further, the negative reinforcement value of a preventive action would be increased among persons who experience both higher levels of susceptibility and intolerance of uncertainty (Carleton, 2016a, Carelton 2016b).

There are few studies investigating the relationship between mindfulness and preventive health behaviors. However, researchers who have examined this link reported that mindful awareness was associated with more preventive health behavior (O’Brien, Horan, et al., 2019). Further, mindfulness-based interventions (acceptance and commitment therapy) was been associated with improved safety behavior and preventive health behaviors (O’Brien, Singh, et al, 2019). The underlying mechanisms between mindfulness and safe behavior are primarily cognitive. Zhang and Wu (2014) discussed how greater awareness to internal and external stimuli, executive attention, and self-regulation of implicit, automatic impulses influence dual process pathways (i.e. automatic and conscious) for safe behavior. The FFMQ-nonreactivity results are in line with this research.

The inverse relationship between FFMQ-nonjudgment and PPE use was unexpected. An examination of the FFMQ-nonjudgment items (e.g., “I think some of my emotions are bad or inappropriate and I shouldn’t be feeling them”) provides insight into how this subscale might be associated with less PPE use. Note that higher level of *disagreement* with these types of items comprise the FFMQ-nonjudgment score. Disagreeing with such items could underlie nonadherence to PPE use. Specifically, one can nonjudgmentally view a thought that nonadherence with a recommended action is “wrong.” As such, unpleasant feelings associated with nonadherence (e.g., guilt, fear of negative social evaluation) would be diminished and made less aversive. This finding represents the double-edged impact of mindfulness in the health domain. Nonjudgment can perhaps be used to both promote acquisition and adherence to preventive health behaviors as well as nonadherence.

Post trauma symptoms and physical symptoms were strongly predicted by susceptibility variables and intolerance of uncertainty. Participants were (and continue to be) living in a social context where frequent and intense threat information was being generated by news media. Exacerbating this exposure to ubiquitous threat were the leadership responses that were frequently at odds with research-supported medical recommendations. There is ample research evidence indicating that threat combined with uncertainty and lack of control creates psychophysiological reactions that can result in many adverse outcomes including reduced immunity (Contrada & Baum, 2011; Taha et al., 2013a; 2013b; Taha, Matheson, Cronin, & Anisman, 2013). Thus, the high levels of PTSD and physical health symptoms in this sample can plausibly be linked not only to the uncertainties of COVID-19 but also intensely threatening media coverage combined with a disorganized leadership response.

The psychological and physical health of persons in the USA as well as adoption of life-saving COVID-19 prevention behaviors can be improved with the introduction of consistent, measured, and research-supported messaging from governmental leadership, public health officials, and media sources. Additionally, accurate susceptibility information combined with research-guided information about the effectiveness of various preventive health behaviors can aid in improving adherence to behaviors that can reduce COVID-19 incidence. Finally, mindfulness-based interventions focused on acceptance and nonreactivity to improve adherence and also reduce psychophysiological distress.

## Limitations

The survey was completed at a single point in time which limits causal inference. We are proposing a temporal sequence where COVID-19 objective risk exposure, demographic and health risk factors, and individual differences predict PPE use, post-trauma symptoms, and physical symptoms. However, the temporal sequence could have a different form. It could be that higher levels of distress, could cause a person to perceive themselves to be more vulnerable, more intolerant of uncertainty, and less mindful. A time series approach would help determine the plausibility of this argument.

A second set of limitations is related to the participant selection process and the use of MTurk workers. In terms of the former, it may be that persons who were experiencing higher levels of distress were more apt to participate in the study. As a result, it cannot be determined how well the descriptive statistics and regression findings generalize to a broader USA population. In relation to the latter limitation, there have been extensive analyses of the representativeness and characteristics of MTurk samples relative to other participant recruitment strategies (see Chandler & Shapiro, 2016). MTurk samples are more representative than college students and convenience samples drawn from small university communities, but less diverse in some ways than national probability samples. However, Chandler and Shapiro (2016) noted that the national probability samples are biased in that they rely on telephone methods which skews their results to older and more conservative participants. Additionally, as was evident in our sample, MTurk workers tend to have higher educational attainment and are more likely to be male. Finally, MTurk workers have been demonstrated to report higher levels of distress relative to other types of samples (Chandler & Shapiro, 2016).

There are strengths to the use of an MTurk sample as well. Collection of data from a heterogenous sample allowed us to be more certain that effects did not pertain to a single geographic location, occupation, or type of participant. Although some research has shown that MTurk participants display higher rates of anxiety and depression, this phenomenon is at least partially combatted through screening for response quality (Ophir et al., 2019). Additionally, this feature of our data may have helped us avoid range restriction in our sample. Finally, although researchers have identified various threats to validity that may be possible in research conducted on a crowdsourced platform, such as subject inattentiveness, demand characteristics, and repeated participation, the present study utilized best practices to mitigate such threats. Specifically, the study utilized attention checks, data screening, and avoiding signaling cues (Cheung et al., 2017).

## Data Availability

Data is available from first author upon request.

